# Rituximab for autoimmune myasthenic syndromes: a retrospective cohort study in myasthenia gravis and Lambert-Eaton myasthenic syndrome

**DOI:** 10.64898/2026.08.18.26360320

**Authors:** Roxanna Chamani Cheri, Ulrike Grittner, Paolo Doksani, Carla Dusemund, Lea Gerischer, Meret Luise Herdick, Sarah Hoffmann, Sophie Lehnerer, Frauke Stascheit, Maike Stein, Andreas Meisel, Philipp Mergenthaler

**Affiliations:** Charité – University Medical Center Berlin, Center for Stroke Research Berlin, Berlin, Germany; Charité – University Medical Center Berlin, Institute of Biometry and Clinical Epidemiology, Berlin, Germany; Charité – University Medical Center Berlin, Department of Neurology with Experimental Neurology, Berlin, Germany; Charité – University Medical Center Berlin, Neuroscience Clinical Research Center, Berlin, Germany; Berlin Institute of Health at Charité – University Medical Center Berlin, Digital Health Center, Berlin, Germany; Radcliffe Department of Medicine, University of Oxford, Oxford, UK

**Author notes:** Correspondence: Philipp Mergenthaler, Charité – University Medical Center Berlin, Center for Stroke Research Berlin, Charitéplatz 1, 10117 Berlin, Germany,.

**Keywords:** myasthenia gravis, Lambert-Eaton myasthenic syndrome, rituximab, B-cell depletion, retrospective cohort study

## Abstract

**INTRODUCTION:** Myasthenia gravis (MG) and Lambert-Eaton myasthenic syndrome (LEMS) are autoimmune diseases of the neuromuscular junction resulting in fatigable muscle weakness. Rituximab (RTX) is used to treat patients refractory to standard immunosuppression, but evidence for its efficacy remains inconsistent. Here, we analyzed real-world data on the clinical course and side effects of RTX in MG and LEMS patients.

**METHODS:** This was a single-center study of all patients diagnosed with MG (n=64) or LEMS (n=5) treated with RTX from 2011 until 2021. Outcomes of RTX treatment were recorded retrospectively with Myasthenia Gravis Foundation of America Post-Intervention Status (MGFA-PIS), number of rescue therapies, myasthenic crises, and steroid dose at 1-year and 2-year follow-ups.

**RESULTS:** MGFA-PIS improved at both 1-year (y) and 2-y follow-up compared with baseline. Incidence rates of rescue therapies per 100 person-months (95% CI) decreased from 15.0 (11.8–18.8) at baseline to 7.5 (4.7–12.3) at 1-y and 4.3 (2.5–7.8) at 2y-follow-up. The number of patients without myasthenic crises within one year increased from baseline (49, 86.0%) to 1y-follow-up (55, 96.5%). Median (IQR) daily steroid dose decreased from 10 (5– 22.5) mg/d at baseline to 4 (0–10) mg/d at 1y-follow-up, and to 2.5 (0–10) mg/d at 2y-follow-up.

**CONCLUSION:** This study indicates that RTX was associated with a stabilized clinical course and decreased steroid use in patients with autoimmune myasthenic syndromes, including those with thymoma-associated MG. Our data suggest that therapeutic benefit is apparent within the first year of treatment and is maintained through two years.

## Introduction

Autoimmune myasthenic syndromes comprise myasthenia gravis (MG) and Lambert-Eaton myasthenic syndrome (LEMS). Both are rare autoimmune disorders of the neuromuscular junction [1]. In MG, autoantibodies are directed against epitopes of the postsynaptic membrane of the neuromuscular junction [1–3], whereas LEMS is characterized by autoantibodies against presynaptic voltage-gated calcium channels [4, 5]. In both, the result is impaired neuromuscular transmission resulting in a characteristic fatigable muscle weakness [3]. LEMS is clinically distinguishable from MG in that muscle weakness typically begins in the proximal lower limbs and frequently involves the autonomic nervous system [6]. Next to symptomatic treatment with acetylcholinesterase inhibitors in MG or 3,4-diaminopyridine for LEMS, long-term immunosuppression is required for most patients. However, an estimated 10-15% of patients with generalized (g)MG do not respond to standard immunosuppressive therapies, such as corticosteroids, azathioprine, or mycophenolate mofetil. These patients are at higher risk of myasthenic crisis (MC) [7–10] and more frequently require rescue therapies (plasmapheresis, immunoadsorption, intravenous immunoglobulins) [11, 12]. Several targeted therapies have recently been evaluated in controlled settings in gMG patients with high disease activity, directed against the neonatal Fc receptor (FcRn) in patients with acetylcholine receptor (AChR)-antibodies or muscle-specific tyrosine kinase (MuSK) antibodies [13–15], or against the complement system in AChR-antibody-positive patients [16–19]. However, patients with other antibody subtypes remain without approved targeted options, and the cost of these therapies is high. Corticosteroids and steroid-sparing immunosuppression are also used for treating LEMS [20], but novel targeted therapies are not approved. Further treatment strategies to achieve sustained symptom control and increase quality of life in patients with refractory gMG [21] and LEMS [22] are needed.

B cell depleting therapies have a long history in the treatment of neurological autoimmune diseases. Rituximab (RTX) is a monoclonal anti-CD20 antibody which depletes B cells and their precursors, but not longer-lived cells of the B cell lineage [23]. Originally developed for B cell lymphoma, RTX has also been found effective in autoimmune diseases, including those of the nervous system [24]. RTX is well tolerated and long-term data in rheumatoid arthritis underline its safety [25]. However, the efficacy of RTX in myasthenic syndromes remains uncertain. While case reports and retrospective studies largely support the use of RTX in severe gMG [26–30] or in patients with MuSK antibodies [31, 32], the recent BeatMG and RINOMAX randomized trials have reached differing conclusions [33, 34]. Notably, the CD19-targeting monoclonal antibody inebilizumab, which depletes B cells and plasma cells [35], has recently been approved for patients with AChR- and MuSK-antibody-positive gMG [36]. For LEMS, the evidence is scarce, comprising only single case reports and one small case series [37–39].

To better understand efficacy and safety of long-term treatment with RTX in gMG and LEMS, we conducted a single-center retrospective observational study. Because both conditions are IgG-mediated disorders of neuromuscular transmission for which B cell depletion offers a common therapeutic rationale, we studied them together as autoimmune myasthenic syndromes. The main objectives were to investigate whether RTX treatment leads to clinical improvement as measured by the Myasthenia Gravis Foundation of America-Post Intervention Status (MGFA-PIS), and to assess changes in rescue therapies, incidence of MC and in steroid doses at 1- and 2-year (y)-follow-up compared to baseline (i.e. first RTX dose).

## Methods

### Study design

This was a single-center, retrospective observational cohort study of all patients with myasthenic syndromes treated with RTX between 2011 and 2021 at the Myasthenia gravis Center of Excellence of the Department of Neurology, Charité - University Medical Center Berlin, a large quaternary care center for MG in Germany, accredited by the German Myasthenia Gravis Association.

### Study cohort, inclusion criteria, collected data

The study design, data collection procedures, and analysis plan outlined below were specified before any data were collected. This study included all patients with gMG or LEMS who received their first dose of RTX at our center between 2011 and 2019. The study period including follow-up data assessment was from the date of the first dose of RTX until December 31, 2021. Data extraction and analysis were conducted from July 2019 until June 2025. We recorded data at baseline and for 1- and 2-year follow-up-intervals. The date of the first ever administration of RTX was defined as the date for “baseline” assessment. Follow-up timepoints were defined by the closest recorded patient visit 12 and 24 months after baseline (±3 months).

RTX-treated patients were eligible if they were older than 18 at the time of inclusion in the analysis. Per local clinical consensus and in accordance with national treatment guidelines [40], patients received 2 doses of 1000 mg RTX within 2 weeks as induction and 1000 mg every 6 months as maintenance. Clinical records indicated that this schedule was adjusted in individual patients depending on the patients’ clinical status and/or levels of CD19- and CD20-positive cells. For this study, the two initial doses of RTX in the beginning of therapy were counted as one RTX cycle. Patients had to be treated with RTX for at least 9 months at 1-year and 21 months at 2-year follow-up to be included in the analysis at follow-up. Data were manually extracted from electronic medical records and were collected in a purpose-designed database using REDCap electronic data capture tools [41] hosted by the Neuroscience Clinical Research Center at Charité – University Medical Center Berlin.

For baseline information the following variables were collected: sex, date of birth, date of diagnosis of MG or LEMS, date of first administration of RTX, detected autoantibodies (for MG: AChR, MuSK, low-density lipoprotein receptor-related protein (LRP) 4, seronegative (for AChR, MuSK, LRP4); for LEMS: voltage gated calcium channel PQ-type (VGCC-PQ) antibodies), thymectomy: histology, thymoma grading (WHO, Masaoka), number of steroid-sparing immunosuppressants prior to RTX, steroid therapy prior to RTX, last daily dose of steroids at start of RTX, number of rescue therapies one year prior to RTX (intravenous immunoglobulins, plasmapheresis, immunoadsorption), number of MC one year prior to start of RTX, MGFA score at start of RTX, MGFA-PIS at start of RTX (compared to MGFA and clinical status at diagnosis, see below), number of RTX cycles and cumulative dose of RTX, complications and side effects associated with RTX administration.

Variables of follow-up information included: number of MC and number of rescue therapies since last administration cycle of RTX (i.e., between BL and 1-year follow-up or between 1-year and 2-year follow-up), MGFA, MGFA-PIS, steroid dose in mg/day, number of RTX cycles, cumulative RTX-dose, complications or side effects after RTX administration (including all adverse effects linked to RTX ranging from immediate side effects after infusion to those between BL and 1-year follow-up and 1-year follow-up and 2-year follow-up), information about thymoma relapse, MG-specific co-medication during study period. MGFA-PIS, MGFA score, number of rescue therapies and MC and steroid dose in mg/day at 1-year and 2-year follow-up were used to assess the disease severity and monitor therapy outcomes of MG and LEMS patients treated with RTX.

### MGFA and MGFA-PIS scores

For Myasthenia Gravis Foundation of America score (MGFA) and Myasthenia Gravis Foundation of America - Post Intervention Status (MGFA-PIS) a period of ± 3 months at 1- and 2-year follow-up was tolerated and for baseline values -3 months were tolerated. MGFA-PIS was determined retrospectively by the first author (R.C.C.), as it was not recorded in the medical records. MGFA scores and physicians’ notes on clinical status and changes in medication dose (e.g., steroids or steroid-sparing immunosuppressants) were used to derive changes in MGFA-PIS. MGFA is routinely documented in our center, but MGFA scores were determined retrospectively based on physicians’ notes if they were missing from the patients’ records. MGFA-PIS at baseline was determined by comparing a patient’s recorded clinical status at baseline to the recorded status at diagnosis of MG or LEMS. MGFA-PIS and MGFA at 1- and 2-year follow-ups refer to baseline. Although the MGFA-PIS was originally designed for MG patients, it was also used to evaluate the clinical status of LEMS patients in this study. Complete Stable Remission (CSR), Pharmacological Remission (PR), Minimal Manifestation (MM-0, MM-1, MM-2, MM-3) and Improved (I) were classified as favorable outcomes in MGFA-PIS. An improvement in MGFA score was defined as a decrease of at least one class and worsening in MGFA as an increase of at least one class.

### Rescue therapies

Treatments with intravenous immunoglobulins, plasmapheresis, and immunoadsorption at any point during the follow-up periods were considered as “rescue therapy”. The number of rescue therapies is summarized without giving separate numbers for each kind of rescue therapy. Multiple cycles of the same rescue therapy given during a single hospital stay were counted as one event. To compare outcomes at baseline with 1- and 2-year follow-up, we counted the number of rescue therapies one year prior to the first dose of RTX. For number of rescue therapies, a period of ±3 months at 1- and 2-year follow-up was tolerated.

### Steroids

A period of ±3 months at 1- and 2-year follow-up was tolerated for the last steroid dose in mg/day. For baseline, the steroid dose at the start of RTX was recorded. If values were not available at this time, the last known steroid dose prior to start of RTX was used.

### Statistical analyses

Data from gMG and LEMS patients were analyzed as one cohort, as both are antibody-mediated disorders of the neuromuscular junction with overlapping clinical features. Only demographic data are presented in separate tables for each disease. For presentation of descriptive data, median and interquartile range (IQR) were used for ordinal scaled data. For comparisons between baseline and 1- or 2-year follow-up data, only patients with available data at the respective follow-up time points were included in the baseline representation. The corresponding number of patients (’n’) is indicated in the figures and tables. The Wilcoxon signed-rank test for related samples was applied for analyzing MGFA-PIS, MGFA score, last steroid dose, and MC incidence at follow-up time points compared to status prior to RTX for unadjusted analyses. Multiple binary logistic regression was performed for MGFA-PIS outcomes (1 = CSR, PR, MM and Improved; 0 = other MGFA-PIS outcomes). Age at first dose of RTX, disease duration, sex, antibody status and MGFA score were considered as confounders and were included as independent variables. For rescue therapy outcomes, incidence rates and 95% confidence intervals per time point were calculated using separate negative binomial regression models for each time point with log-transformed individual observation time as offset (Figure 3). Incidence rate ratios and 95% confidence intervals for therapy with RTX were calculated using a negative binomial mixed model with log-transformed individual observation time as offset, and a random intercept for individuals (Figure 3, Supplemental Table 8). One model was calculated using only baseline and 1-year follow-up measures, another model using all available measures at baseline, 1- and 2-year follow-ups. For calculating negative binomial mixed models R package lme4 [42], and for the other negative binomial regression models R package MASS [43] was used. SPSS Statistics 26.0.0 or R 4.1.1 (www.r-project.org) were used to perform all other statistical analyses. Microsoft Excel 365 was used to create plots for Figs. 2 and 3 and Graphpad Prism 9 to create plots for Fig. 4. A two-sided significance level of 0.05 was used. This was an exploratory study without adjustment for multiple testing. Therefore, while we report p-values for completeness, they have to be interpreted cautiously.

## Results

### Cohort characteristics

All patients with MG or LEMS treated at our center as of Dec. 31, 2021 (n=1,695) were screened for eligibility for this study (Fig. 1). The study population consisted of 69 patients meeting the eligibility criteria (n=64 MG, Table 1; n=5 LEMS, Table 2) from 71 gMG/LEMS patients treated with RTX at our center (Fig. 1). For 57 patients (n=52 gMG, n=5 LEMS) 1y-follow-up data and for 42 patients (n=38 gMG, n=4 LEMS) 2y-follow-up data were available. Three patients were excluded from follow-up analyses: two received RTX for less than 9 months during the study period, and one patient was switched from RTX to bortezomib and subsequently eculizumab within three months after RTX initiation (Fig. 1). None of these patients reached the pre-specified timepoints for follow-up assessment. Two patients (n=1 gMG, n=1 LEMS) died during the period of observation (see “safety” below). Other reasons for missing 1y-follow-up and 2y-follow-up information are shown in Fig. 1. The median duration of RTX therapy for patients with available 2y-follow-up data (n=42) was 3.5 years (IQR: 2.6-5.2 years, min/max: 2.1/10.3 years). However, all analyses were limited to the observation period of two years after start of RTX (±3 months). Eleven patients (16.0%) used further steroid-sparing immunosuppressants as co-medication during the study period (Supplemental Table 1).

**Fig. 1.**
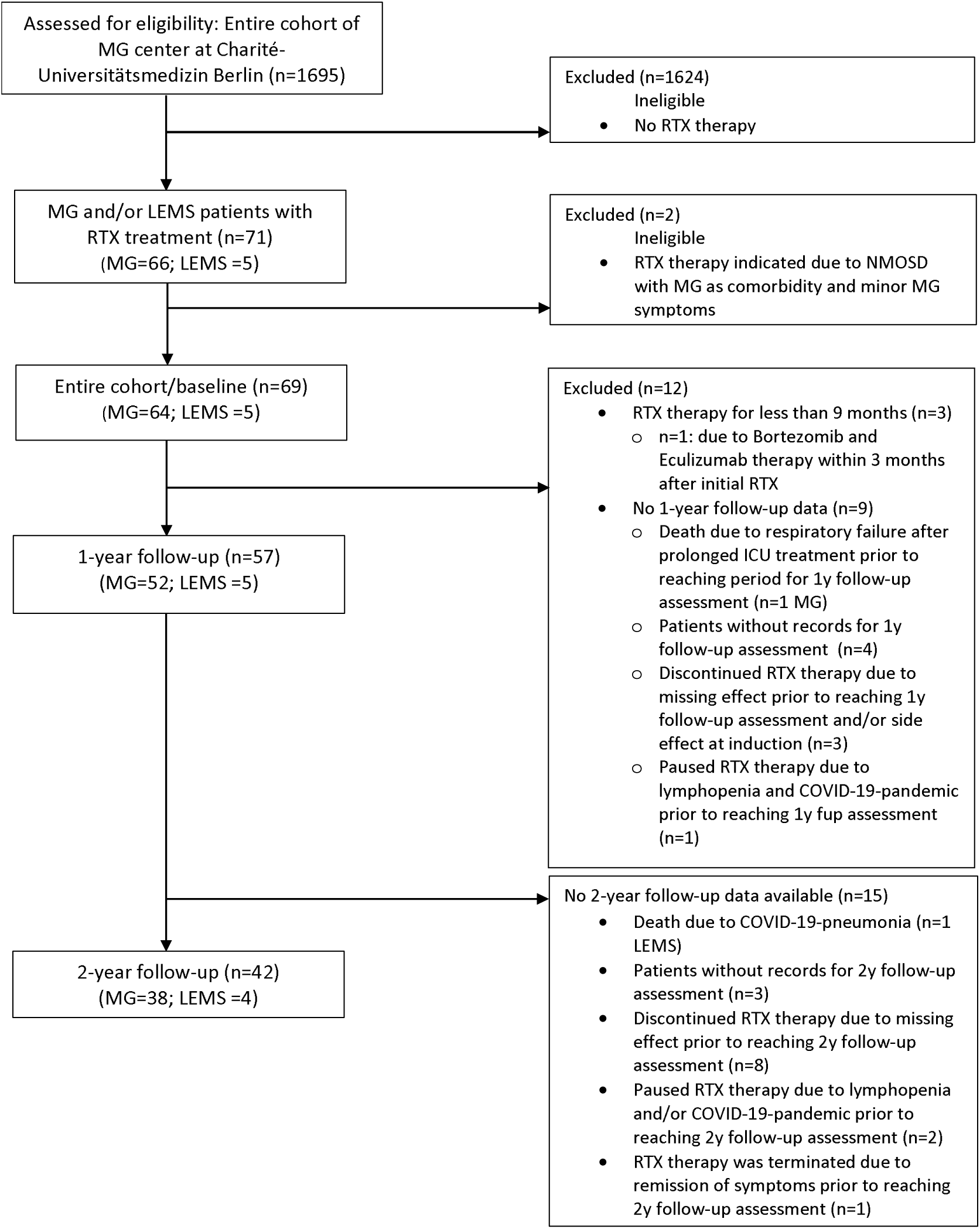
Flow chart about the process of patient enrollment *ICU* intensive care unit, *LEMS* Lambert-Eaton myasthenic syndrome, *MG* Myasthenia gravis, *NMOSD* Neuromyelitis optica spectrum disease, *RTX* Rituximab

**Table 1:**
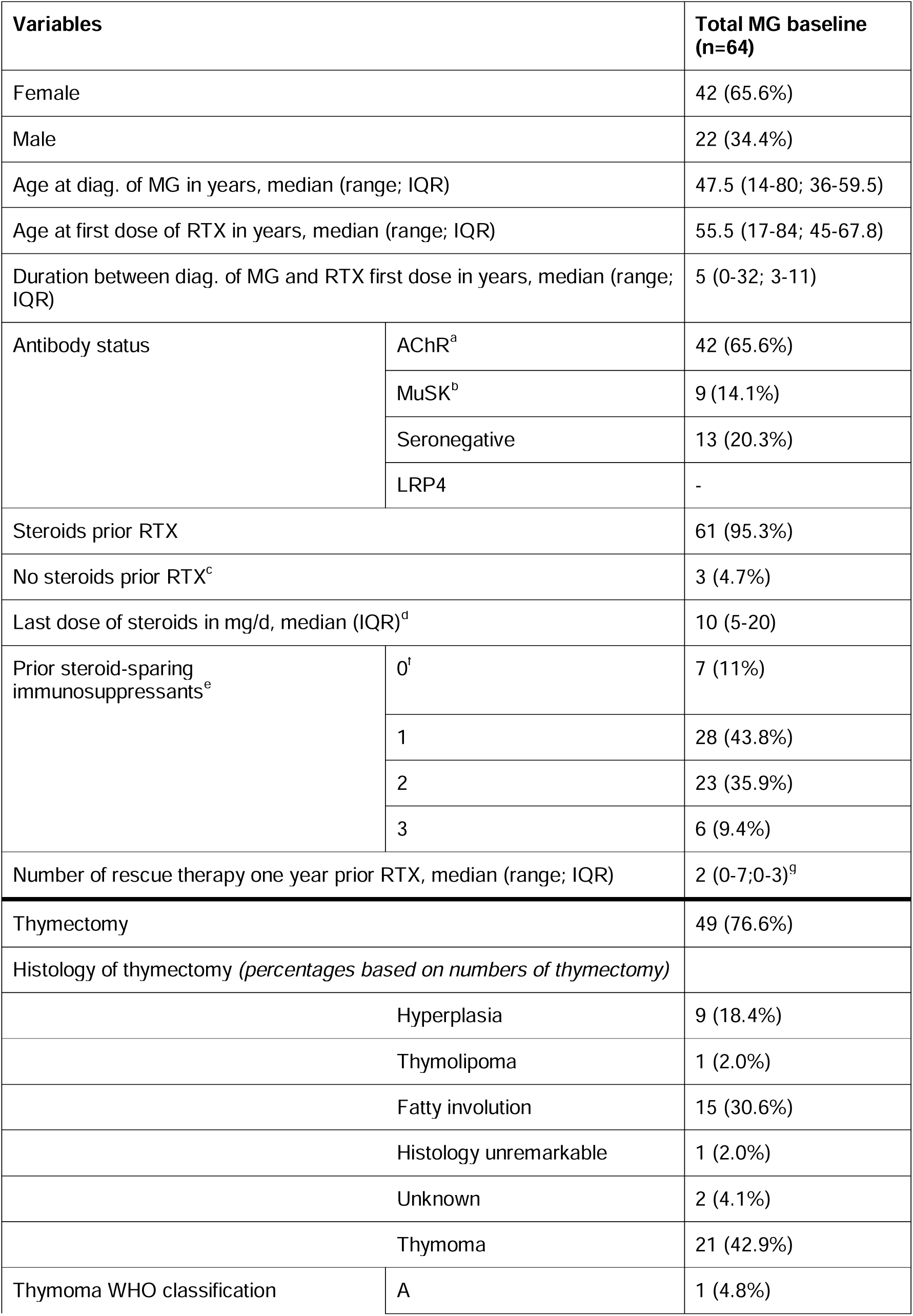
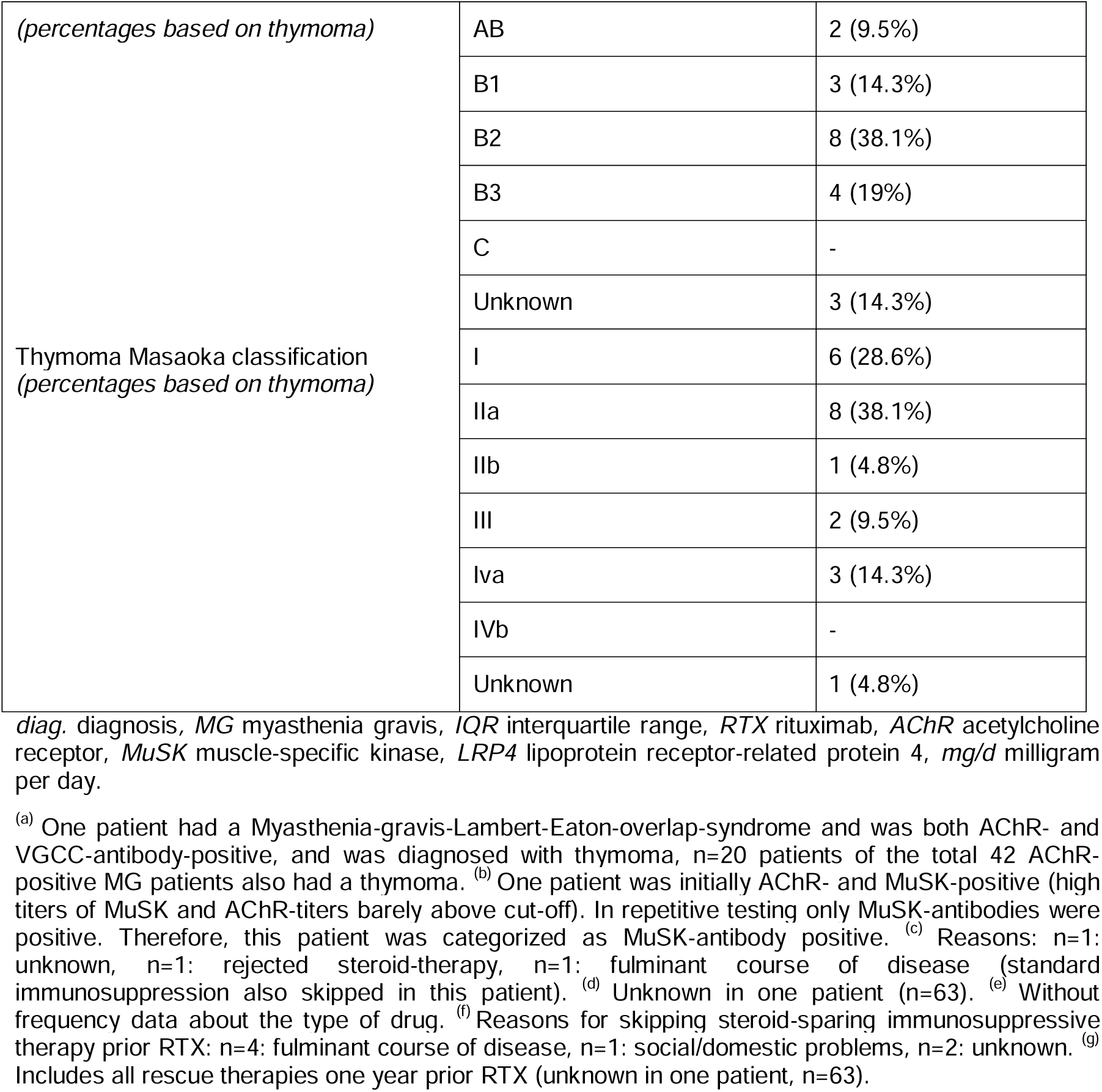
Demographics of Myasthenia gravis patients before start of RTX therapy (baseline)

**Table 2.** Demographics of Lambert-Eaton syndrome patients before start of RTX therapy.

| Variables |  | total LEMS baseline (n=5) |
| --- | --- | --- |
| Female |  | 2 (40%) |
| Male |  | 3 (60%) |
| Age at diagnosis of LEMS in years, median (range, IQR) |  | 63 (31-68; 44.5-67.5) |
| Age at first dose of RTX in years, median (range, IQR) |  | 73 (31-76; 45.5-75) |
| Duration between diag. of LEMS and RTX first dose in years, median (range; IQR) |  | 5 (0-13; 1-10) |
| Etiology | Autoimmune | 5 (100%) |
|  | Paraneoplastic | - |
| Antibody status | VGCC-PQ | 4 (80%) |
|  | Seronegative | 1 (20%) |
| Prior steroid-sparing immunosuppressants<sup>a</sup> | 0 | 2 (40%) |
|  | 1 | 2 (40%) |
|  | 2 | 1 (20%) |
| Steroids prior RTX |  | 5 (100%) |
| Last dose of steroids in mg/d, median (range; IQR) |  | 5 (0-100; 2-55) |
| Number of rescue therapy one year prior RTX, median (range; IQR)<sup>b</sup> |  | 2 (0-3; 0.5-3) |

*VGCC-PQ* PQ-type voltage-gated calcium channel, *mg/d* milligram per day.
<sup>(a)</sup> Without frequency data about the type of drug. <sup>(b)</sup> Includes all rescue therapies one year prior RTX.

### Clinical characteristics and treatment prior to RTX for MG patients

Forty-two gMG patients were female (65.6%, Table 1). Median age (range; IQR) at diagnosis of gMG was 48 years (14-80; 36-60) and at first dose of RTX 56 years (17-84; 45-68). Median duration (range; IQR) between diagnosis of gMG and first dose of RTX was 5 years (0-32; 3-11). AChR-antibodies were present in 42 (65.6%) patients, 9 (14.1%) patients had MuSK-antibodies, and 13 (20.3%) patients were seronegative for AChR, MuSK, and LRP4. Most patients received steroids before the initiation of RTX (n=61; 95.3%). One patient had a MG-LEMS-overlap-syndrome and was both AChR- and VGCC-antibody-positive and was diagnosed with thymoma (Table 1). Forty-nine patients (76.6%) underwent thymectomy, of which 21 patients (42.9% of thymectomized, 32.8% of MG) were diagnosed with thymoma (Table 1). Steroid-sparing immunosuppressants, such as azathioprine, mycophenolate mofetil, or methotrexate had been used by 57 (89.1%) patients prior to RTX. Reasons for not taking steroids or steroid-sparing immunosuppressants are given in Table 1. The median (IQR) of the last steroid dose prior to RTX was 10 (5-20) mg/day (Supplemental Table 2). The median (range; IQR) number of rescue therapies one year prior to initiation of RTX was 2 (0-7; 0-3; Table 1). Forty-nine patients underwent thymectomy, of which 21 (42.9%) patients had a thymoma (Table1).

### Clinical characteristics and treatment prior to RTX for LEMS patients

Five LEMS patients (two [40%] female) were included in our study cohort (Table 2). Median (range; IQR) age at diagnosis of LEMS was 63 years (31-68; 44.5-67.5), and at first dose of RTX was 73 years (31-76; 45.5-75). Median duration (range; IQR) between diagnosis of LEMS and first dose of RTX was 5 (0-13; 1-10) years. VGCC-PQ-antibodies were present in four (80%) patients, and one patient (20%) was seronegative. Etiology in all LEMS patients was autoimmune rather than paraneoplastic. Three (60%) patients received steroid-sparing immunosuppressants before initiation of RTX. All patients had received steroids (n=5; 100%) with a median (range; IQR) last steroid dose prior to RTX of 5 (0-100; 2-55) mg/day. The median (range; IQR) number of rescue therapies one year prior to initiation of RTX was 2 (0-3; 0.5-3, Table 2).

### Safety and tolerability of RTX in the entire study population

Sixteen of 69 patients (23.2%) developed one or more side effects due to RTX infusions (Supplemental Table 3). These were allergic reactions (n=2; 2.9%), direct infusion reactions (n=4; 5.8%), and further transient symptoms (n=10; 14.5%). One multi-morbid 82-year-old male with MuSK-antibody-positive gMG (1.4% of 69 patients; Fig. 1) died 6.5 months after first administration of RTX after prolonged intensive care treatment for six months due to respiratory failure, having been unable to wean from invasive mechanical ventilation. This patient had presented with an MC within one month after initial administration of RTX and had received the second dose of the RTX induction cycle as part of the treatment for the MC. The ventilation situation was complicated by ventilator-induced lung injury on the basis of underlying chronic obstructive pulmonary disease, ventilator-associated pneumonia due to Klebsiella and Pseudomonas, peripheral mucus retention with a combined restrictive-obstructive ventilatory disorder, pulmonary arterial hypertension, and fibrotic changes of the pulmonary architecture. B cells were depleted at the time ventilator-associated pneumonia was diagnosed and were not reassessed thereafter. One LEMS patient (1.4% of 69 patients) died because of SARS-CoV-2-associated pneumonia during the observation period at the end of 2020 after receiving four cycles of RTX (cumulative dose 5000 mg, last dose ∼5 months before death, COVID-19 vaccination status and peripheral B cell count at time of death unknown) and did not reach the pre-specified window of observation for 2y-follow-up.

### Clinical outcomes in the entire study population

In patients with 1y-follow-up data available, 27 of 57 patients (47.4%) showed a favorable outcome in MGFA-PIS (“Improved” or MM≤) with an improvement in the distribution of MGFA-PIS outcomes at 1y-follow-up compared to baseline (Fig. 2, Supplemental Table 4). Patients with 2y-follow-up data available (n=42) reached a favorable outcome in 52.4% (n=22) at 1y-follow-up, and in 45.2% (n=19) at 2y-follow-up. The distribution of MGFA-PIS outcomes at 1y-follow-up or 2y-follow-up was improved compared to baseline. There was no difference of MGFA-PIS outcome distribution between 1y-follow-up and 2y-follow-up (Fig. 2, Supplemental Table 4). One patient died due to a myasthenic crisis outside of the pre-specified intervals and was therefore not included in the statistical analysis (see above). Binary logistic regression revealed no associations between tested variables and “improved” or better MGFA-PIS outcomes (Supplemental Table 5).

**Fig. 2.**
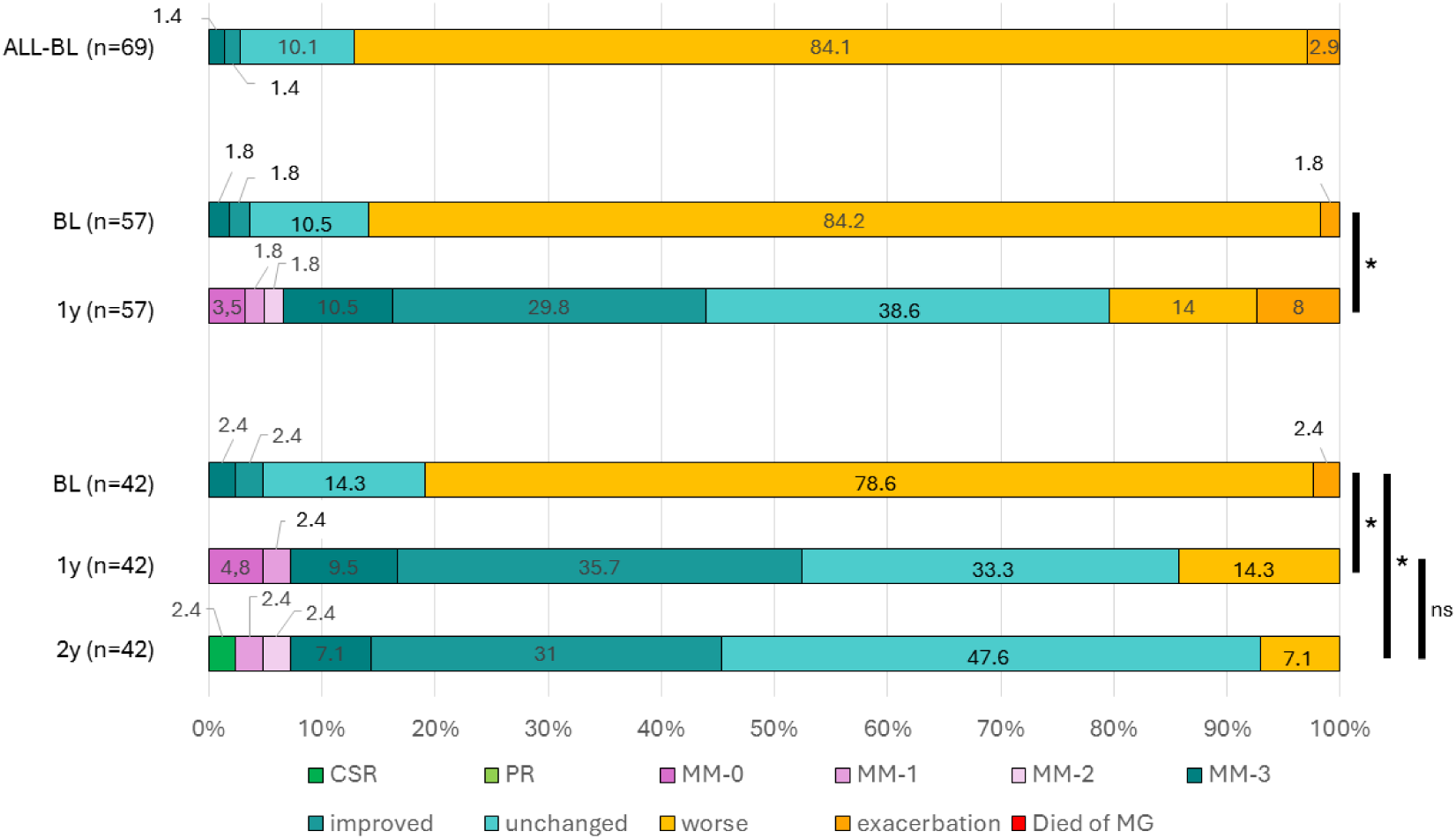
Graphical overview of MGFA-post intervention status (PIS) outcomes as percentage of the respective total patient number stratified by follow-up period. Shown are representations of the entire cohort (ALL-BL, n=69), all patients with 1 year follow-up data at baseline (BL, n=57) and at 1-year follow-up (1y, n=57), and all patients with 2-year follow-up data at baseline (BL, n=42), 1 year follow-up (1y, n=42), and 2 year follow-up (2y, n=42). Note: all items for MGFA PIS are shown in the legend underneath the graph to indicate full assessment. BL is defined as first administration of RTX. At baseline, MGFA-PIS reflects clinical status at first RTX administration compared with status at diagnosis of MG or LEMS. See Supplemental Table 4 for complete numerical values. *Statistical testing: Wilcoxon regression on distribution of MGFA-PIS, *p<0.01, ns - not significant. CSR -Complete Stable Remission, PR - Pharmacological Remission, MM - Minimal Manifestation*.

Similar changes were observed in the MGFA score, which is widely used as a pragmatic parameter for clinical assessment (Supplemental Table 6, Supplemental Table 7). Distribution of MGFA scores for patients with 1y-follow-up (n=57) or 2y-follow-up (n=42) at 1- and 2-y-follow-ups were improved compared to baseline (Supplemental Table 7).

### Use of rescue therapy and incidence of myasthenic crisis

The fraction of patients without need for rescue therapies increased from baseline (29.4%, 20 of 68, one patient with unknown data at baseline) to 1y-follow-up (61.4%, 35 of 57) and 2y-follow-up (61.9%, 26 of 42), respectively (Fig. 3; Supplemental Table 8). Importantly, incidence rates per 100 person months (95% CI) for the number of rescue therapies in the 12 months prior to the observation date decreased from baseline (15.0 (11.8-18.8)) to 1y-follow-up (7.5 (4.7-12.3)) and further at 2y-follow-up (4.3 (2.5-7.8)). The incidence rate ratios at 1y-follow-up (0.48; 95%CI: 0.35-0.67; p<0.001) and at 2y-follow-up (0.34; 95%CI: 0.26-0.45; p<0.001) indicate that the number of therapies after starting RTX was substantially reduced by approximately 50% and more than 60%, respectively, compared to the time before start of RTX (Fig. 3; Supplemental Table 8).

**Fig. 3.**
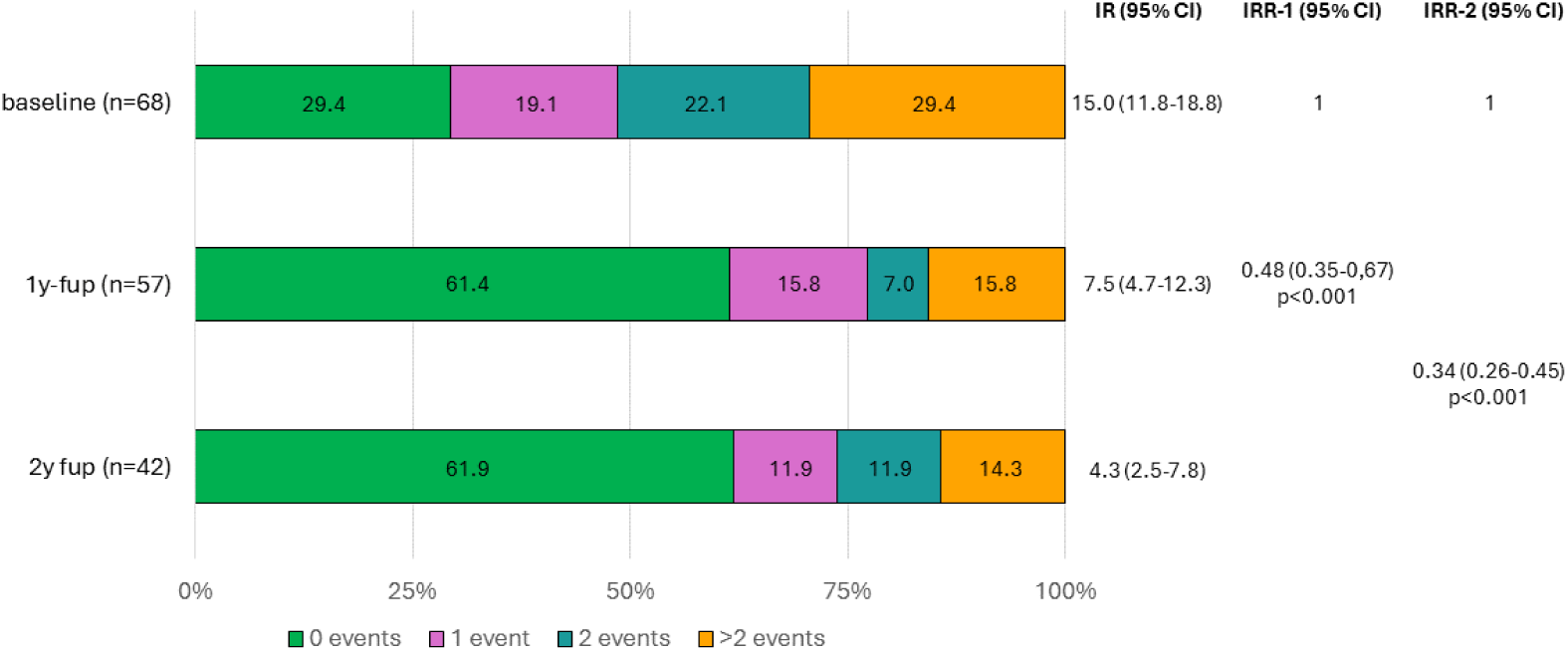
Graphical overview of the need for rescue therapies as percentage of the respective group. Also shown are the incidence rate per 100 person months (IR), the incidence rate ratio (IRR) for RTX therapy including only baseline and 1y-follow-up measures (IRR-1), and the IRR for RTX therapy including all available measures. Baseline was defined as the 12-month period before the start of RTX therapy. Note: data for one patient was unknown at baseline (N=68). *See Supplemental Table 8 for complete numerical values*.

The percentage of patients with no MC increased from 86.0% (49 of 57) in the year prior to RTX induction (i.e., baseline) to 96.5% (55 of 57) at 1y-follow-up (Table 3). At the 2y-follow-up 92.9% (39 of 42) of the patients did not have an MC in the preceding period of observation.

**Table 3:**
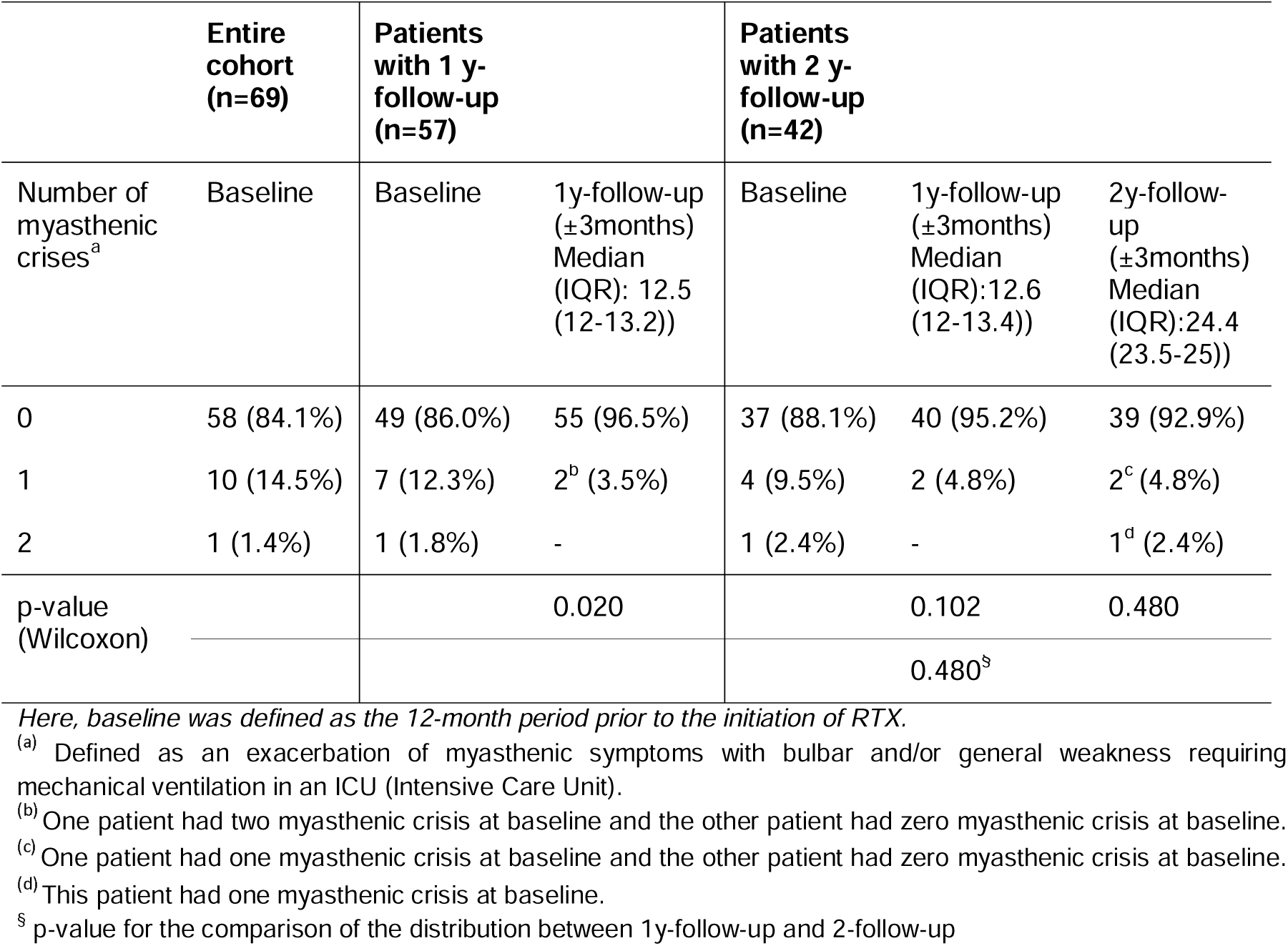
Number of myasthenic crises.

### Steroid use after RTX

The steroid dose decreased during the observation period for most patients compared to baseline across all antibody classes (Fig. 4, supplemental table 2). However, there was no further decrease of steroid doses between 1y-follow-up and 2y-follow-up (Fig. 4B, Supplemental table 2). A decrease in steroid dose was seen in 61.4% (n=35/57, patients with only 1y-follow-up data) of patients at 1y-follow-up (Supplemental Table 9) with a median (IQR) of 4 (0-10) mg/d compared to baseline (10 (5-22.5) mg/d; Supplemental Table 2). Patients with 2y-follow-up (n=42) also showed a decrease of their steroid dose at 1y-follow-up (n=30/42; 71.4%) with a median (IQR) of 4 (0-10) mg/d, and at 2y-follow-up (n=29/42; 69.1%) with median (IQR) of 2.5 (0-10) mg/d compared to baseline (10 (5-26.3) mg/d; Supplemental Table 2 which also shows the mean for these doses; Supplemental Table 9). The steroid dose remained unchanged in nearly half of patients at 2y-follow-up compared to the same patients at 1y-follow-up (n=20/42; 47.6%; Supplemental Table 9).

**Fig. 4.**
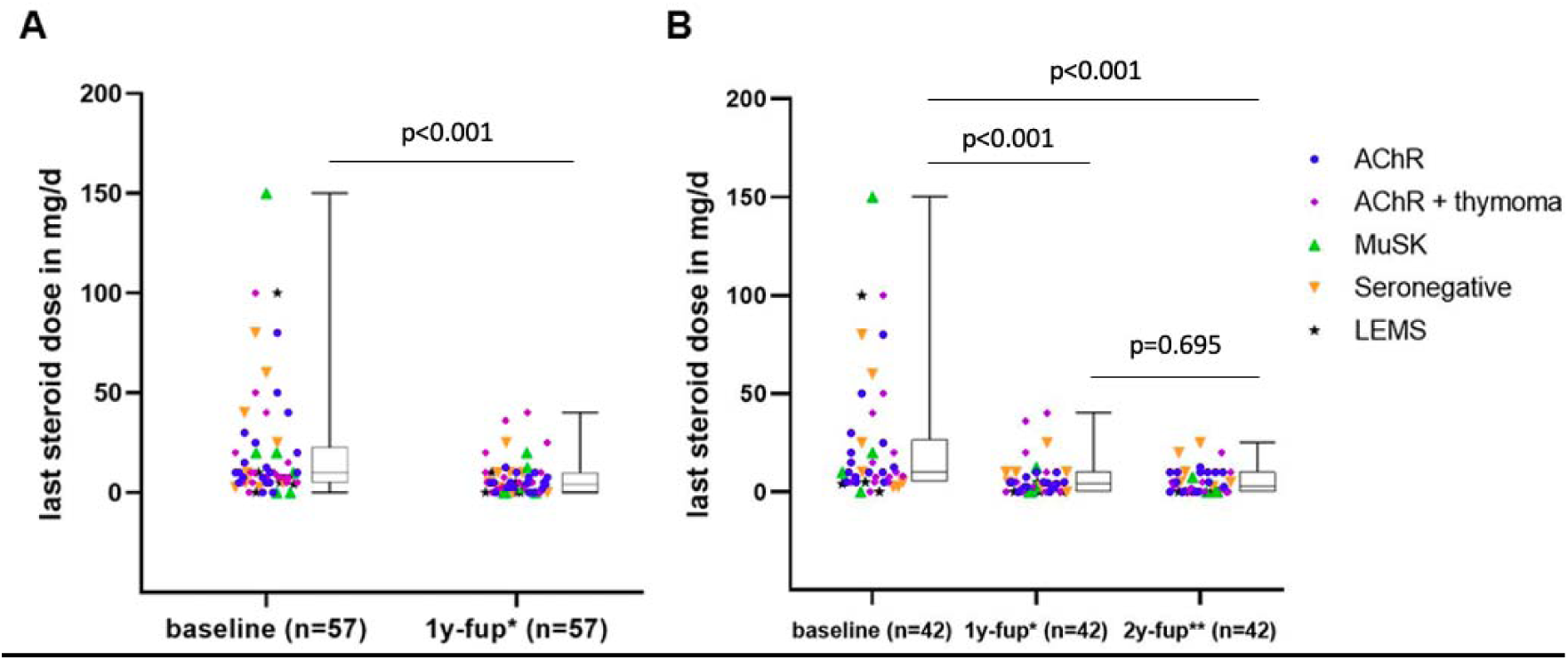
**A** Last recorded steroid doses at baseline and 1-year follow-up (*1y-follow-up)*, * time period of 1y-follow-up in months (Median (IQR)): 12.5 (12-13.3)) for patients with recorded 1-year follow-up (n=57). **B** Steroid doses at baseline, 1- and 2-year follow up only for patients with recorded 2-year follow-up (n=42). *time period of 1y-follow-up in months (Median (IQR)): 12.6 (11.8-13.4), **time period of 2y-follow-up in months (Median (IQR)): 24.5 (22.3-25).

The percentage of patients with steroid doses below the Cushing threshold (≤7.5 mg/d) and with 0 mg increased during the observation period (Supplemental Table 10). More than half of the patients had steroid doses below the Cushing threshold at 1y-follow-up (68.4%) and 2y-follow-up (71.4%). Eighteen patients were not treated with steroids when RTX was started, of which 2 patients were naïve to steroids prior to RTX and for 1 patient the status was unknown (Table 1).

### RTX cycles and cumulative dose of RTX

At 1y-follow-up the median (IQR) number of RTX cycles was 3 (2-3) with a cumulative RTX median (IQR) dose of 4000 (3000-4000) mg (Supplemental Table 11). At 2y-follow-up the median (IQR) number of RTX cycles was 4 (3-5) with a cumulative median (IQR) dose of 5000 (4000-6000) mg (Supplemental Table 11).

## Discussion

In this retrospective observational study, we investigated the clinical efficacy and safety of RTX treatment in patients with autoimmune myasthenic syndromes irrespective of autoantibody status. Our data show clinical improvement within one year of initiating RTX, together with a reduced need for rescue therapies and lower steroid doses. Myasthenic crises occurred less frequently after RTX initiation than in the year before. These findings corroborate clinical experience and published case series, and contribute to a better understanding of the effect of RTX therapy on the clinical course of myasthenic syndromes. Our cohort also included one patient with MG-LEMS overlap syndrome, positive for both AChR and VGCC antibodies, and with thymoma. Notably, 21 of 64 MG patients (32.8%) had thymoma-associated MG (TAMG), to our knowledge the largest such series treated with RTX reported to date. Since TAMG is associated with more severe disease and a less favorable prognosis than non-thymomatous MG [44], the overall benefit observed in our cohort was achieved in a population enriched for difficult-to-treat MG.

Previously, prospective unblinded studies, retrospective studies, and case reports have investigated the efficacy of RTX in myasthenic syndromes [27–29, 45–51]. Altogether, these studies showed improvement in clinical scores, decreased need for rescue therapies and decreased steroid doses with RTX therapy, and that RTX was generally well-tolerated. Further, some case reports and one retrospective study found a positive effect of RTX on the clinical status of LEMS patients [37–39]. Our findings are consistent with these observations. However, a randomized, double-blind, placebo-controlled phase 2 study of RTX in AChR antibody-positive gMG (BeatMG) reached its futility endpoint for the primary composite outcome of steroid reduction with clinical stability, indicating that a larger trial would be unlikely to demonstrate the predefined benefit [33]. Yet, the sample size of BeatMG was fixed by the limited availability of RTX rather than derived from a power calculation, so the futility threshold was set against the number of patients available rather than a prespecified clinically meaningful effect [33]. A smaller but clinically meaningful benefit could therefore have been missed. We observed a decrease in the use of steroids for 61.4% of patients at 1y-follow-up, which is similar to the observations for the study and control groups of the BeatMG trial [33]. However, the retrospective and observational nature of our study does not allow inference about how steroid use would have changed in a control group without RTX. A retrospective analysis of AChR antibody-positive gMG patients compared RTX treatment in new-onset and refractory gMG with conventional immunosuppression [30]. The study found that early treatment with RTX led to remission more rapidly and more frequently than in refractory gMG or in patients treated with conventional immunosuppression [30]. The need for rescue therapies (i.e., IVIg, plasma exchange, high-dose intravenous steroids) was substantially reduced in patients receiving RTX compared to standard immunotherapy [30].

Likewise, the recent prospective randomized RINOMAX trial led by the same group found that RTX treatment in new-onset gMG was linked to low steroid doses [34]. Patients were more likely to reach minimal disease manifestation at 16 weeks without the need for rescue therapies when treated with RTX compared to placebo [34]. In these two randomized trials, RTX produced only modest changes in the secondary outcomes of Myasthenia Gravis Activities of Daily Living Scale (MG-ADL) and quantitative myasthenia gravis (QMG) scores, in contrast to the marked benefit reported in observational cohorts. This discrepancy has been attributed both to differences in trial populations and design and to the limitations inherent in uncontrolled data [52]. Together with our data, these findings support a therapeutic role for RTX in autoimmune myasthenic syndromes, beyond MuSK-antibody-positive gMG.

Patients with AChR antibody-positive gMG often show a favorable response to standard immunosuppressive therapies, and, therefore, data about the use of RTX in these patients are scarce [53]. A systematic review of 13 studies on the use of RTX in AChR-positive gMG found clinical improvement and reduction of the average prednisone dose as well as the number of rescue therapies in most patients of this MG subgroup [53]. In contrast, the BeatMG did not support a steroid-sparing benefit in mild-to-moderately symptomatic AChR-gMG [33]. Nevertheless, the trial reported a decrease of MG relapses demanding rescue therapy in patients treated with RTX versus the placebo group, indicating a stabilizing effect of RTX in generalized AChR-MG [33].

More modern B cell-targeting therapies may be equally or more effective than RTX for the treatment of myasthenic syndromes [54]. Recently, the MINT trial of the CD19-positive B cell-depleting monoclonal antibody Inebilizumab reported improved function and reduced disease severity in AChR- or MuSK-antibody-positive gMG [36] which has led to subsequent approval of a B cell-depleting therapy for gMG in several countries. There are no head-to-head trials of B cell depletion with FcRn or C5 inhibitors, but a recent network meta-analysis suggested that FcRn inhibitors, C5 inhibitors, and CD19-targeted B cell depletion have comparable efficacy [52]. Furthermore, the apparent inferiority of RTX in these comparisons might in part reflect differences in trial populations, since the estimated efficacy of RTX increased once age and sex were accounted for [52].

CD20-targeting antibodies (RTX) deplete naïve and memory B cells but spare CD20-negative plasmablasts and long-lived plasma cells [23, 55], whereas anti-CD19 antibodies (inebilizumab) target a broader range of the B lineage, and proteasome inhibitors (e.g., bortezomib) act directly on antibody-secreting cells as shown in an experimental model of MG [56]. RTX is more effective in MuSK-than in AChR-antibody-positive disease [57], consistent with the origin of MuSK IgG4 autoantibodies in short-lived, CD20-derived plasmablasts [55]. Whether broad depletion of the B cell lineage confers clinical benefit over narrower depletion of CD20-positive cells remains mechanistically untested. The only interventional trial of bortezomib in MG was terminated for recruitment failure without reporting efficacy outcomes [58, 59], leaving case reports of bortezomib alone [60, 61] or in combination with RTX [62]. The anti-CD19 and anti-CD20 trials differ too much in design and population to permit mechanistic inference [52]. However, after the approval of inebilizumab it is unlikely that there will be another prospective trial for RTX or its inclusion in prospective head-to-head trials. Consequently, real-world observational data, such as ours, may help characterize the therapeutic value of RTX in autoimmune myasthenic syndromes.

A small percentage of our study population (n=11; 16%) was treated with steroid-sparing immunosuppressants during the study period, suggesting a more severe course of MG. Of these, 55% (n=6/11) did not need steroid-sparing immunosuppressants at 1y-follow-up anymore. Therefore, RTX might be most effective in severely affected patients, as has been suggested previously [48, 63]. Consistent with this, we found a reduction in the need for rescue therapies as well as a reduction of the incidence of MC after induction of RTX. In the absence of a control group, comparing similar timeframes before and after the induction of RTX therapy in the same patients served as an internal control. Our results therefore suggest that the clinical endpoints “need for rescue therapy” and “incidence of MC” or an MGFA-PIS of “Improved” or better (“Improved” / “MM” / “CSR” / “PR”) could be suitable outcome parameters to investigate the efficacy of RTX or other B cell depleting therapies for treating myasthenic syndromes in future prospective trials.

National [40] and international guidelines as well as systematic reviews and expert consensus [53, 57, 64] favor the use of RTX for gMG in certain circumstances, particularly for patients presenting with MuSK antibodies. We did not find MuSK-positive antibody status to be an independent baseline variable predicting clinical efficacy (measured by MGFA-PIS of “Improved” or better) using logistic regression. This finding is not in line with prior studies, which show that response to RTX therapy is greater in gMG patients with MuSK-antibodies than in patients with AChR-antibodies [27, 31, 32, 46, 65]. Our contrasting finding might be due to the small sample size of our cohort, the lack of a control group, a selection bias since all patients treated with RTX at our center were included in this study, or due to the study design.

Our autoantibody-agnostic analysis indicates that clinical benefit is apparent within one year of starting RTX in most patients, and is unlikely to emerge thereafter. Thus, pragmatically, MG patients who have not improved on RTX in that timeframe might then be switched to other advanced (and much more expensive) therapies. In this context, a recent retrospective study found the C5 inhibitor eculizumab to be more effective than RTX in improving muscle strength as measured by the QMG score [66]. However, the risk for MC remained the same with eculizumab and RTX treatment [66]. Our data, as well as prospective data [34], suggest that the need for rescue therapies and the frequency of MCs decrease with RTX therapy. Our data further support a pragmatic use of RTX in myasthenic syndromes irrespective of the antibody subtype, and clinical evaluation of the therapeutic effect after one year of treatment. Accordingly, RTX has been used in the treatment of lipoprotein receptor-related protein (LRP)4-antibody-positive MG [67].

In our study, RTX was well-tolerated with side effects mainly restricted to infusion reactions, which is in line with other observations [27, 33, 34, 53, 57] and a recent network meta-analysis [52]. However, one MuSK-MG patient died in association with the initial administration of RTX, and it is possible that the ventilator-associated pneumonia that exacerbated the clinical course in this patient was related to B cell depletion. One LEMS patient died of SARS-CoV-2-associated pneumonia during the period of observation early in SARS-CoV-2 pandemic (see “safety” above). Our data are insufficient to derive a risk for death after SARS-CoV-2 infection related to RTX treatment. However, recent studies found an association between RTX treatment and poor outcome or death in MG patients after COVID-19 [68, 69]. Even though we did not observe cases of progressive multifocal leukoencephalopathy (PML) in our study population, two cases of PML have been reported in MG patients treated with RTX [26, 70].

Limitations of our study arise from its pooled, retrospective, and single-center design. MG and LEMS were analyzed as a single cohort of autoimmune myasthenic syndromes, reflecting their shared status as IgG-mediated disorders of neuromuscular transmission with broadly common treatment approaches and a shared rationale for B cell depletion. The two conditions nevertheless differ in target antigen and in the site of the transmission defect, and with five LEMS patients the cohort was too small to permit separate analysis. Conclusions specific to LEMS should therefore be drawn with caution. MGFA-PIS assessments and MGFA scores (where missing from the source data) were determined retrospectively from patient records by a single rater without independent verification or blinding to timepoint, which could lead to overestimation of favorable effects. Even though guidelines favor the use of RTX [40], it has never been approved for gMG. Our study population may therefore be biased to severe courses of the disease, or patients who had been refractory to other therapies, selected for off-label therapy prior to the approval of FcRn or C5 inhibitors. According to pre-specified criteria of our study protocol, we investigated outcome at 1y- and 2y-follow-up timepoints after first dose of RTX. The pre-specified follow-up timepoints and observation windows (see Methods) meant that patients who discontinued RTX or died before reaching these intervals were excluded from the respective analyses, which may have enriched the cohort for treatment responders. Immunoglobulin levels and infection episodes were not systematically recorded, so we cannot report on hypogammaglobulinemia or infection risk, which are among the principal long-term safety concerns with repeated RTX administration. Finally, it cannot be ruled out that patients received rescue therapies or were treated for MC at other centers, and that not all such events were accounted for in the patients’ records.

In conclusion, this study indicates that RTX may be an efficacious therapy with few side effects in patients with MG and LEMS, beyond its use in new-onset generalized MG patients [30, 34]. Our data and studies [27, 30, 34] support expecting therapeutic efficacy within one year after treatment initiation. Future randomized controlled trials may consider studying clinical endpoints such as “need for rescue therapies” and “MC incidence”, as well as patient reported outcomes, rather than merely steroid use, and may be necessary to further investigate the efficacy of RTX in MG subgroups and in LEMS. Furthermore, future studies should focus on biomarker discovery to identify patients at risk for a critical disease course [10], and to enable proper and early [34] patient stratification for the various novel targeted immunotherapies emerging for myasthenic syndromes.

## Statements and Declarations

### Ethics approval

This study was approved by the ethics committee of Charité - University Medical Center Berlin (EA1/260/19). As per the ethics approval, local regulations, and applicable law, informed consent was neither obtained nor required for this retrospective study. The study was conducted in accordance with the Declaration of Helsinki, the German Federal Data Protection Act, and the EU General Data Protection Regulation, and is reported in accordance with the STROBE statement for observational studies.

### Data availability statement

Ethical approval currently does not permit sharing of raw data. Approval will be sought by the corresponding author upon reasonable request with scientific rationale and sound methodology. Requests for data sharing will be managed in accordance with data access and sharing policies of Charité – University Medical Center Berlin.

## Author contributions

R.C.C. collected, curated, analyzed, discussed and interpreted the data, and wrote the initial draft and edited manuscript. U.G. performed and supervised statistical analyses, and discussed and interpreted the data. P.D., C.D., L.G., M.H., S.H., S.L., F.S., M.S. discussed the data and edited the manuscript for intellectual content. A.M. discussed and interpreted the data, and edited the manuscript for intellectual content. P.M. conceived and supervised all aspects of the study, discussed and interpreted the data, and revised and edited the manuscript. All authors contributed to the submitted version of the paper and approved it for publication.

## Acknowledgements

We thank M. Olszewska for REDCap support, C. Heibutzki, D. Remstedt, N. Baro for patient and study management, and S. Lischewski, S. Märschenz and M. Heinold for administrative support.

