## Supplemental Information for "Rituximab for autoimmune myasthenic syndromes: a retrospective cohort study in myasthenia gravis and Lambert-Eaton myasthenic syndrome"

**Short title:** Rituximab in myasthenia gravis and LEMS


Roxanna Chamani Cheri[1], Ulrike Grittner[2], Paolo Doksani[3,4], Carla Dusemund[3,4], Lea Gerischer[3,4], Meret Luise Herdick[3,4], Sarah Hoffmann[3,4], Sophie Lehnerer[3,4,5], Frauke Stascheit[3,4], Maike Stein[3,4,5], Andreas Meisel[1,3,4], Philipp Mergenthaler[1,3,6]

[1] Charité – University Medical Center Berlin, Center for Stroke Research Berlin, Berlin, Germany

[2] Charité – University Medical Center Berlin, Institute of Biometry and Clinical Epidemiology, Berlin, Germany

[3] Charité – University Medical Center Berlin, Department of Neurology with Experimental Neurology, Berlin, Germany

[4] Charité – University Medical Center Berlin, Neuroscience Clinical Research Center, Berlin, Germany

[5] Berlin Institute of Health at Charité – University Medical Center Berlin, Digital Health Center, Berlin, Germany

[6] Radcliffe Department of Medicine, University of Oxford, Oxford, UK

### Supplemental Tables

**Supplemental Table 1:** Comedication during study period

| RECORD-ID | baseline | 1y-follow-up | 2y-follow-up |
|---|---|---|---|
| Pat-01[a] | AZA (25mg 1-0-0)<br>Predni (5mg 1-0-0) | AZA (75 mg 1-0-0)<br>Predni (40 mg 1-0-0)<br>(11.4 months) | AZA (50 mg 1-0-0)<br>Predni (10 mg 1-0-0)<br>(27 months) |
| Pat-02 | AZA (50mg 1,5-0-1,5)<br>Predni (15 mg 1-0-0) | AZA : /<br>Predni (2.5mg 1-0-0)<br>(11,7 months) | AZA : /<br>Predni: /<br>(23.6 months) |
| Pat-03 | AZA (50 mg 1-0-0)<br>Predni: / | AZA: /<br>Predni: /<br>(13.3 months) | AZA: /<br>Predni: /<br>(22.1 months) |
| Pat-04 | AZA (50 mg 1-0-1)<br>Predni 60/5mg alternate | AZA 50mg (1-0-1)<br>Predni: /<br>(11.8 months) | AZA: 75mg (1-0-0)<br>Predni: /<br>(20 months) |
| Pat-05 | MMF (500mg 3-0-2)<br>Predni:/ | MMF: /<br>Predni: /<br>(14.7 months) | MMF:/<br>Predni: /<br>(20.7 months) |
| Pat-06 | MMF (500mg 2-0-2)<br>Predni (10 mg 1-0-0) | MMF (500mg 1-0-1)<br>Predni (25mg 1-0-0)<br>(13 months) | no data available |
| Pat-07 | AZA (25mg 1-0-0)<br>Predni: (40mg 1-0-0) | AZA (25mg 1-0-0)<br>Predni: (2.5 mg 1-0-0)<br>(13months) | no data available |
| Pat-08 | MMF (500mg 2-0-0-2)<br>Predni: / (5mg 1-0-0) | MMF:/<br>Predni: (5 mg 1.5-0-0)<br>(12.5 months) | no data available |
| Pat-09 | MTX 15mg (1x/week)<br>Predni: / | MTX:/<br>Predni: /<br>(13.1 months) | no data available |
| Pat-10 | AZA: (50mg 2-0-2)<br>Predni: (10mg 1-0-0) | AZA: /<br>Predni: (10mg 1-0-0)<br>(12.4 months) | no data available |
| Pat-11 | AZA (50mg 4-0-0)<br>Predni (10 mg 1.5-0-0) | no data available | no data available |

*AZA* azathioprine*, MMF* Mycophenolate mofetil; Predni prednisolone.
[(a)] This patient was also treated with azathioprine because of autoimmune hepatitis.

**Supplemental Table 2:** Steroid dose during study period

| **Last steroid dose in mg/d** | Baseline (n=57) | 1y-follow-up (n=57) | Baseline (n=42) | 1y-follow-up (n=42) | 2y-follow-up (n=42) |
|---|---|---|---|---|---|
| Median (IQR) | 10 (5-22.5) | 4 (0-10) | 10 (5 -26.3) | 4 (0-10) | 2.5 (0-10) |
| Mean (SD) | 21.1(29.7) | 6.4(8.6) | 24.6(33.2) | 6.4 (9) | 4.9 (6.2) |

*mg/d:* milligram per day*, IQR:* interquartile range, *SD:* standard deviation

**Supplemental Table 3:** Side effects of RTX infusion

| Side effects of RTX infusion | | Number of patients |
|---|---|---|
| Allergic reactions | | 2 (2.9%) |
| Direct infusion reactions[a] | | 4 (5.8%) |
| | - Nausea | 4 (100%) |
| | - Vomiting | 2 (50%) |
| | - Hypotension | 1 (25%) |
| | - Fever | 1 (25%) |
| | - Diarrhea | 1 (25%) |
| Transient symptoms | | 10 (14.5%) |
| | - General weakness | 2 (2.9%) |
| | - Insomnia | 1 (1.4 %) |
| | - Dizziness | 1 (1.4 %) |
| | - Burning sensation of the throat | 1 (1.4 %) |
| | - Eczema on the scalp | 1 (1.4 %) |
| | - Thrombophlebitis | 1 (1.4 %) |
| | - Intertriginous erythema | 1 (1.4 %) |
| | - Persistent burning of the tongue area[b] | 1 (1.4 %) |
| | - Persistent pain in the forearm[c] | 1 (1.4 %) |

(a) Some patients developed more than one direct infusion reaction. (b) Of otherwise inexplicable cause and persisting during the entire period of observation. (c) After first RTX infusion, which was the cause for rejection of further RTX infusions; percentages referring to total baseline (n=69).

**Supplemental Table 4:** MGFA-PIS outcomes

| | **Entire cohort (n=69)** | **Patients with 1y-follow-up (n=57)** | | **Patients with 2y-follow-up (n=42)** | | |
|---|---|---|---|---|---|---|
| MGFA-PIS: categories | Baseline | Baseline | 1y-follow-up (±3months) Median (IQR): 12.5 (12.1-13.5)) | Baseline | 1y-follow-up (±3months) Median (IQR):12.6 (12-13.8)) | 2y-follow-up (±3months) Median (IQR):24.3 (22.1-24.7)) |
| CSR | - | - | - | - | - | 1 (2.4%) |
| PR | - | - | - | - | - | - |
| MM-0 | - | - | 2 (3.5%) | - | 2 (4.8%) | - |
| MM-1 | - | - | 1 (1.8%) | - | 1 (2.4%) | 1 (2.4%) |
| MM-2 | - | - | 1 (1.8%) | - | - | 1 (2.4%) |
| MM-3 | 1 (1.4%) | 1 (1.8%) | 6 (10.5%) | 1 (2.4%) | 4 (9.5%) | 3 (7.1%) |
| Improved | 1 (1.4%) | 1 (1.8%) | 17 (29.8%) | 1 (2.4%) | 15 (35.7%) | 13 (31%) |
| Unchanged | 7 (10.1%) | 6 (10.5%) | 22 (38.6%) | 6 (14.3%) | 14 (33.3%) | 20 (47.6%) |
| Worse | 58 (84.1%) | 48 (84.2%) | 8 (14%) | 33 (78.6%) | 6 (14.3%) | 3 (7.1%) |
| Exacerbation | 2 (2.9%) | 1 (1.8%) | - | 1 (2.4%) | - | - |
| Died of MG[a] | - | - | - | - | - | - |
| p-value (Wilcoxon) | - | | <0.001 *compared to 1y-follow-up baseline* | | <0.001 *compared to 2y-follow-up baseline* | <0.001 *compared to 2y-follow-up baseline* |
| | | | | | 0.874[§] | |

*Note: At baseline, MGFA-PIS reflects clinical status at first RTX administration compared with status at diagnosis of MG or LEMS.*

*MGFA-PIS* Myasthenia Gravis Foundation of America- Post-intervention Status, *CSR* Complete Stable Remission, *PR* Pharmacological Remission, *MM* Minimal Manifestation, *MG* Myasthenia Gravis.

[a] One patient died due to a myasthenic crisis during the period of observation. The patient did not reach the pre-specified interval for inclusion in follow-up and is therefore not reported in this Table. See Results ("Safety") for details. [§]There was no difference in the distribution of MGFA-PIS of patients with 2y-follow-up at 2y-follow-up compared to 1y-follow-up (p=0.874).

**Supplemental Table 5:** Binary logistic regression for variables association with MGFA-PIS outcomes of “Improved” or better

| | **1y-follow-up (n=57)** | | | **1y-follow-up of 2y-follow-up (n=42)** | | | **2y-follow-up (n=42)** | | |
|---|---|---|---|---|---|---|---|---|---|
| Independent variables | Odds ratio | 95% CI | p-value | Odds ratio | 95% CI | p-value | Odds ratio | 95% CI | p-value |
| Age at start of RTX in years | 0.97 | 0.93-1.01 | 0.093 | 0.97 | 0.93-1.01 | 0.180 | 0.96 | 0.92-1.01 | 0.109 |
| Disease duration in years | 1.01 | 0.93-1.09 | 0.904 | 0.97 | 0.87-1.09 | 0.602 | 1.00 | 0.89-1.18 | 0.992 |
| Sex (male/female) | 1.38 | 0.43-4.44 | 0.592 | 1.01 | 0.29-4.51 | 0.985 | 1.00 | 0.21-4.48 | 0.998 |
| MuSK-antibody status | 0.49 | 0.07-3.56 | 0.481 | 0.33 | 0.02-4.32 | 0.379 | 0.11 | 0.00-1.85 | 0.125 |
| AChR-antibody status | 0.72 | 0.21-2.54 | 0.613 | 0.51 | 0.11-2.41 | 0.392 | 0.265 | 0.04-1.44 | 0.125 |
| MGFA clinical classification[a] | 1.44 | 0.443-4.661 | 0.546 | 2.95 | 0.72-12.11 | 0.134 | 1.736 | 0.42-7.10 | 0.443 |
| Constant[b] | 7.90 | - | 0.145 | 14.30 | - | 0.164 | 54.19 | - | 0.060 |

[(a)] At baseline, MGFA clinical classification outcomes were divided in two groups: MGFA 0/I/II and MGFA III/IV/V. [(b)] Constant: model intercept, reported for completeness. It does not provide clinical interpretation.

**Supplemental Table 6:** Change in MGFA score

| MGFA score at | **1y-follow-up (±3 months) (n=57)** | **1 y-follow-up (±3 months) (n=42)** | **2y-follow-up (±3 months) (n=42)** | **2y-follow-up (±3 months) (n=42)** |
|---|---|---|---|---|
| Compared to | Baseline (n=57) | Baseline (n=42) | | 1y-follow-up of patients with 2y-follow-up (n= 42) |
| improved | 20 (35.1%) | 18 (42.9%) | 20 (47.6%) | 8 (19%) |
| unchanged | 35 (61.4%) | 22 (52.4%) | 19 (45.2%) | 28 (66.7%) |
| worsened | 2 (3.5%) | 2 (4.8%) | 3 (7.1%) | 6 (14.3%) |

*MGFA* Myasthenia Gravis Foundation of America

**Supplemental Table 7:** Distribution of MGFA scores

| | **Entire cohort (n=69)** | **Patients with 1 y-follow-up (n=57)** | | **Patients with 2 y-follow-up (n=42)** | | |
|---|---|---|---|---|---|---|
| MGFA scores | Baseline | Baseline | 1 y-follow-up (±3months) Median (IQR): 12.6 (11.9-13.6)) | Baseline | 1 y-follow-up (±3months) (Median (IQR):12.7 (11.9-13.5)) | 2 y-follow-up (±3months) (Median (IQR): 24.4 (23.7-25.5)) |
| 0 | - | - | 3 (5.3%) | - | 2 (4.8%) | 3 (7.1%) |
| I | 1 (1.4%) | 1 (1.8%) | 1 (1.8%) | 1(2.4%) | 1 (2.4%) | 3 (7.1%) |
| II | 28 (40.6%) | 25 (43.9%) | 35 (61.4%) | 16 (38.1%) | 26 (61.9%) | 23 (54.8%) |
| III | 31(44.9%) | 26 (45.6%) | 17 (29.8%) | 21 (50%) | 13 (31%) | 13 (31%) |
| IV | 7 (10.1%) | 4 (7%) | 1 (1.8%) | 3 (7.1%) | - | - |
| V | 2 (2.9%) | 1 (1.8%) | - | 1 (2.4%) | - | - |
| p-value (Wilcoxon, distribution of score values) | | | <0.001 (compared to 1y-follow-up baseline) | | <0.001 (compared to 2y-follow-up baseline) | <0.001 (compared to 2y-follow-up baseline) |
| | | | | | 0.412[§] | |

*MGFA* Myasthenia Gravis Foundation of America.

§ No difference in the distribution of MGFA scores of patients with 2y-follow-up at 2y-follow-up compared to 1y-follow-up (p=0.412).

**Supplemental Table 8:** Rescue therapies after RTX

| | **Baseline**[a] (n=68) | **1y-follow-up** (n=57) (±3months) (Median (IQR):12.5 (12.0-13.10)) | **2y-follow-up** (n=42) (±3months) (Median (IQR):24.4 (23.4-24.9)) |
|---|---|---|---|
| Number of patients with…, n (%) | | | |
| 0 events | 20 (29.4%) | 35 (61.4%) | 26 (61.9%) |
| 1 | 13 (19.1%) | 9 (15.8%) | 5 (11.9%) |
| 2 | 15 (22.1%) | 4 (7.0%) | 5 (11.9%) |
| >2 events | 20 (29.4%) | 9 (15.8%) | 6 (14.3%) |
| Incidence rate per 100 person months (95% CI) | 15.0 (11.8-18.8) | 7.5 (4.7-12.3) | 4.3 (2.5-7.8) |
| Incidence rate ratio (95% CI; p-value) for RTX therapy, n=68 patients, 125 measures (only including baseline and 1y- follow-up measures) | 1 (reference) | 0.48 (0.35-0.67; <0.001) | |
| Incidence rate ratio (95% CI; p-value) for RTX therapy, n=68 patients, 167 measures (including all available measures) | 1 (reference) | 0.34 (0.26-0.45; <0.001) | |

*IRR* Incidence Rate Ratio.

[a] 12 months before start of RTX therapy; data for one patient was unknown at baseline.

**Supplemental Table 9:** Changes in steroid use (in number of patients)

| | **1y-follow-up (±3 months) (n=57)** | **1y-follow-up (±3 months) (n=42)** | **2y-follow-up (±3 months) (n=42)** | **2y-follow-up (±3 months) (n=42)** |
|---|---|---|---|---|
| Compared to | Baseline (n=57) | Baseline (n=42) | | 1y-follow-up of patients with 2y-follow-up (n=42) |
| decreased | 35 (61.4%) | 30 (71.4%) | 29 (69.1%) | 12 (28.6%) |
| unchanged | 18 (31.6%) | 9 (21.4%) | 8 (19%) | 20 (47.6%) |
| increased | 4 (7%) | 3 (7.1%) | 5 (11.9%) | 10 (23.8%) |

**Supplemental Table 10:** Percentages of steroid doses below the Cushing threshold (≤7.5 mg/d) and at 0 mg

| Last Steroid dose | baseline (n=68[a]) | 1y-follow-up (n=57) | 2y-follow-up (n=42) |
|---|---|---|---|
| below Cushing threshold[b] | 22 (32.4%) | 39 (68.4%) | 30 (71.4%) |
| at 0 mg | 6 (8.8%) | 17 (29.8%) | 17 (40.5%) |

(a) Unknown in one patient. (b) 7.5 mg per day for prednisone and prednisolone; patients with 0 mg of steroids are included in numbers of last steroid dose < cushing threshold dose

**Supplemental Table 11:** RTX cycles, cumulative dose of RTX

| Variables | 1y-follow-up (n=57) (±3 months) | 2y-follow-up (n=42) (±3 months) |
|---|---|---|
| Number of RTX-cycles, Median (IQR)[a] | 3 (2-3) | 4 (3-5) |
| Cumulative dose of RTX in mg, Median (IQR) | 4000 (3000-4000) | 5000 (4000-6000) |

*RTX* Rituximab, *IQR* interquartile range, *mg* milligram.

[(a)] First administration includes two administrations with a two-week interval of rituximab with 1000 mg each and is counted as one RTX cycle.
